# Risk of evolutionary escape from neutralizing antibodies targeting SARS-CoV-2 spike protein

**DOI:** 10.1101/2020.11.17.20233726

**Authors:** Debra Van Egeren, Alexander Novokhodko, Madison Stoddard, Uyen Tran, Bruce Zetter, Michael Rogers, Bradley L. Pentelute, Jonathan M. Carlson, Mark Hixon, Diane Joseph-McCarthy, Arijit Chakravarty

## Abstract

As many prophylactics targeting SARS-CoV-2 are aimed at the spike protein receptor-binding domain (RBD), we examined the risk of immune evasion from previously published RBD-targeting neutralizing antibodies (nAbs). Epitopes for RBD-targeting nAbs overlap one another substantially and can give rise to escape mutants with ACE2 affinities comparable to wild type that still infect cells *in vitro*. We used evolutionary modeling to predict the frequency of immune escape before and after the widespread presence of nAbs due to vaccines, passive immunization or natural immunity. Our modeling suggests that SARS-CoV-2 mutants with one or two mildly deleterious mutations are expected to exist in high numbers due to neutral genetic variation, and consequently resistance to single or double antibody combinations can develop quickly under positive selection.

**One Sentence Summary:** SARS-CoV-2 will evolve quickly to evade widely deployed spike RBD-targeting monoclonal antibodies, requiring combinations that rely on at least three antibodies targeting distinct epitopes to suppress viral immune evasion.

## Introduction

The deployment of vaccines against SARS-CoV-2 brings the question of mutational escape from antibody prophylaxis to the forefront. SARS-CoV-2 is commonly considered to acquire mutations more slowly than other RNA viruses (*1, 2*). However, the SARS-CoV-2 mutation burden and evolutionary rate (1×10^−3^ substitutions per base per year (*2*)) have only been estimated under conditions that favor neutral genetic drift (distinct from antigenic drift) (*3*), in the absence of strong positive selection pressure provided by population-level immunity or other interventions that select for resistance mutations. In immunologically naïve COVID-19 patients, viral load and transmission (*4*) peak near the time of symptom onset, while the host antibody response peaks approximately 10 days later (*5*). Thus, most transmission occurs well in advance of the appearance of a robust humoral response, suggesting limited intrahost immune evasion prior to transmission, consistent with direct genetic evidence from deep sequencing showing little to no positive selection (*6*). Hence, the current evolutionary rate (based primarily on neutral genetic drift) may underestimate the evolutionary potential of the virus to evade nAbs deployed as active immunity (vaccines) or passive immunity (nAb prophylactics). When nAbs are broadly present in the population, population-level selection for antibody-evading, infection-competent viral mutants may result in a rapid resurgence of SARS-CoV-2 infections.

Mutation rates alone offer a limited picture of the ability of viruses to generate successful escape mutations. While some vaccine-preventable viruses have very low mutation rates (such as smallpox, ∼1 × 10^−6^ sub/nuc/yr) (*7*), others have high mutation rates (such as poliovirus, 1 × 10^−2^ sub/nuc/yr) (Table S1). There is a sharp contrast between the high antigenic evolvability of viruses such as influenza (*8*), notable for their evolutionary capacity for immune evasion, and the low antigenic evolvability of viruses like poliovirus, which have proven highly tractable to antibody-mediated prophylaxis via vaccines (*9*) despite a high evolutionary rate (Table S1). Studies of other infectious diseases support the concept that natural selection promotes antigenic evolvability (*10*).

To better understand the potential for immune evasion mediated by SARS-CoV-2 RBD mutations in the presence of nAbs, singly or in combination, we focused on three questions. First, what is the evolutionary cost of harboring nAb-evading RBD mutations? Second, given this evolutionary cost, how likely is it that SARS-CoV-2 patients will harbor viruses with pre-existing nAb-evading RBD mutations as their dominant viral sequence? Third, how rapidly will such nAb-evading RBD mutants become fixed in the population, once nAb vaccines and therapies are deployed widely?

## Results

### There is a low evolutionary cost to developing resistance to RBD-targeting nAbs

To explore the diversity of the B-cell response against the RBD, we catalogued the reported spike RBD epitopes recognized by the natural human immune response. Consistent with prior work (*11*), we found that the reported epitopes show substantial overlap (Fig. 1A-B). Clustering revealed three clusters representing distinct immunogenic sites on the RBD, the largest of which overlaps substantially with the ACE2 binding interface (Fig. 1A, C). These clusters resemble those reported by other groups (*12*). Notably, there was limited evidence for glycosylation in these epitope clusters (Fig. S1). The observed overlap in residues included in epitopes from independently-generated natural human antibodies shows that parts of the RBD surface are repeatedly targeted by the human B-cell response in different individuals. Spontaneous mutations at these key epitope residues could render many nAbs ineffective.

**Figure 1.**
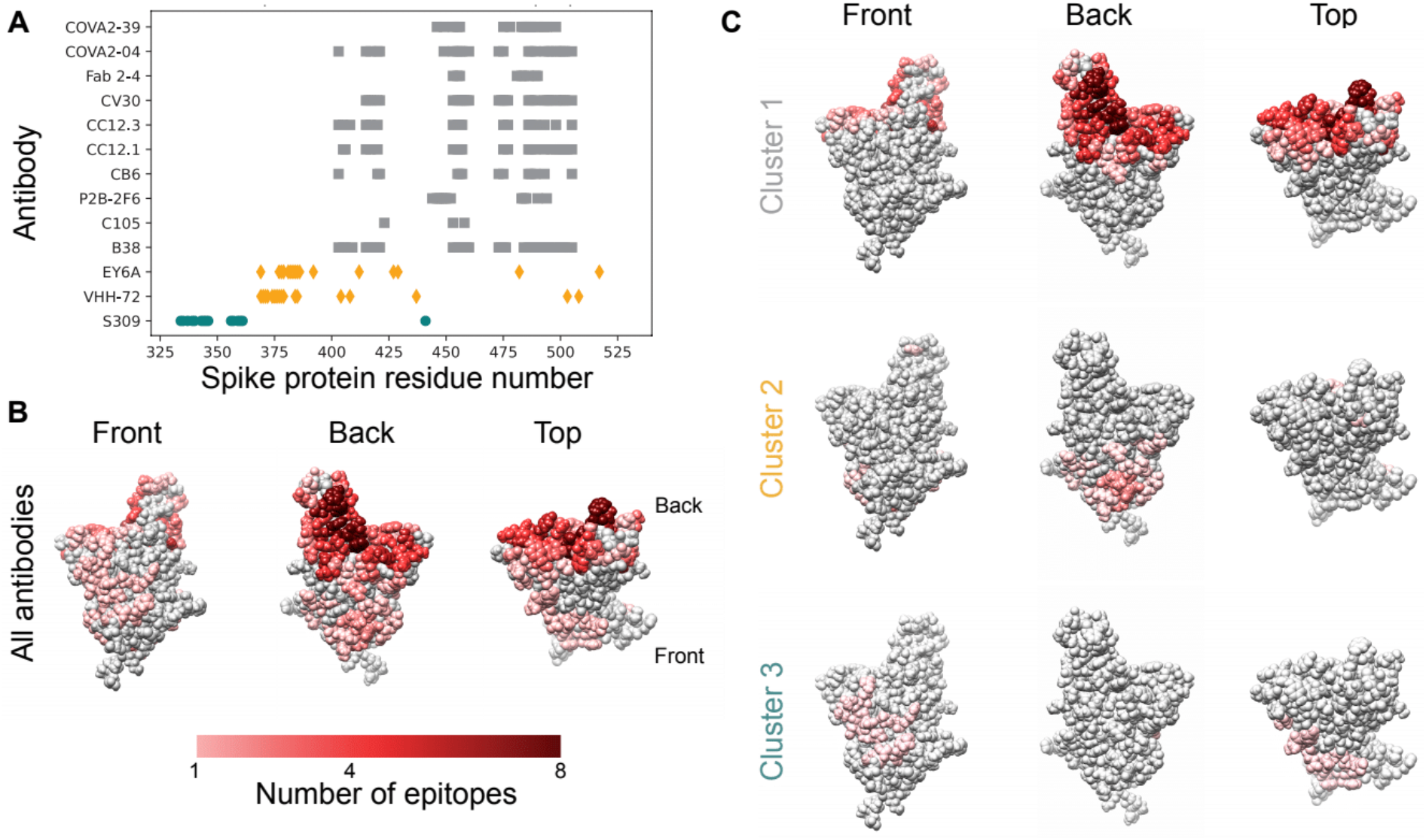
Epitopes for antibodies targeting the spike protein RBD overlap substantially. **A**. Contact residues for spike protein RBD antibody epitopes. Colors and symbols denote antibody clusters: grey squares: cluster 1, yellow diamonds: cluster 2, green circles: cluster 3. **B**. RBD structure with each residue colored by the number of antibody epitopes including it, compiled from PDB data. **C**. RBD structure, colored by the number of antibody epitopes that each residue is part of, by epitope cluster.

Genomic sequencing of SARS-CoV-2 from infected individuals has revealed several point mutations in the RBD, some of which have been shown experimentally to confer resistance to nAbs. As of 8/18/20, multiple amino acid changes have been reported in the GISAID sequence database (*13*) in RBD residues within antibody epitopes (Fig. 2A), showing that SARS-CoV-2 antibody binding region variants are capable of causing human infection. Some of these naturally-occurring variants confer *in vitro* resistance to SARS-CoV-2 nAbs (Fig. 2B) (*14–16*). Additionally, 23 of these experimentally-identified escape mutations (out of 32) have been reported in the GISAID database, many of which do not compromise spike-ACE2 binding when examined *in vitro* (Fig. 2C). This suggests that escape mutants that evade nAb binding have a low evolutionary cost (*17*). In fact, many antibodies have escape mutants that have increased ACE2 binding affinities (Table S2).

**Figure 2.**
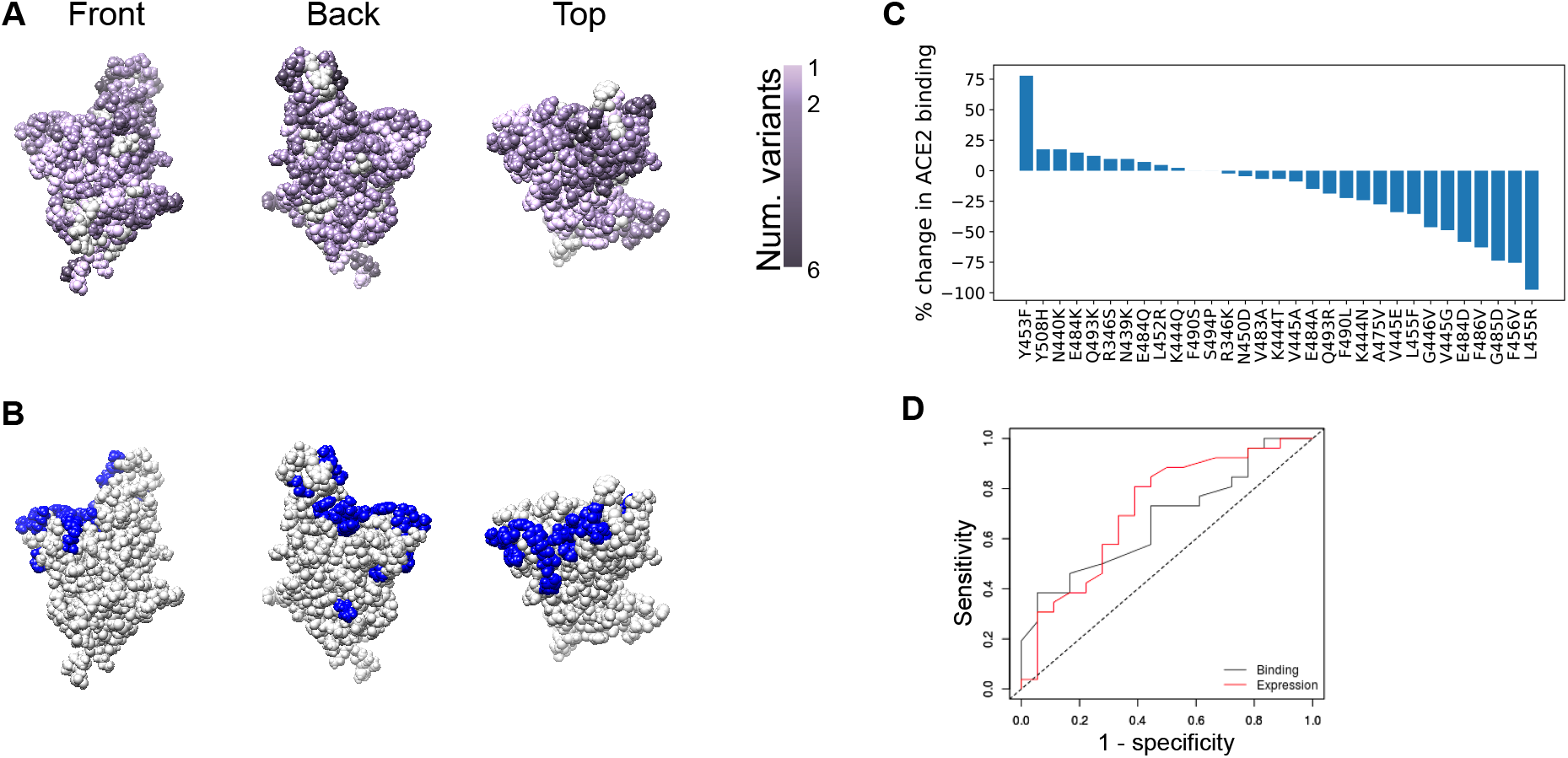
The spike protein RBD tolerates mutations that confer resistance to one or more nAbs. **A**. Spike protein RBD structure, with each residue colored by the number of distinct amino acid changes present in the GISAID sequencing database. **B**. RBD structure with residues at which mutations have been shown to confer escape from antibody neutralization marked in blue. **C**. Experimentally-measured effects of immune escape mutations on ACE2 binding, as taken from (*17*). **D**. ROC curve showing the low predictive value of ACE2 binding measurements (grey) and expression (red) for *in vitro* infectivity of SARS-CoV-2 mutants. Area under the curve (AUC) is 0.67 for ACE2 binding as a predictor of infectivity and 0.72 for RBD expression as a predictor of infectivity.

To further understand the evolutionary cost of escape mutations, we evaluated the link between ACE2 binding affinity and/or RBD expression and viral infectivity (Methods). We determined how well changes in RBD binding to ACE2 or RBD expression caused by a mutation (*17*) predicted a 10% loss of infectivity as measured by luciferase reporter pseudoviral assay (*15*). The low-to-moderate level of sensitivity and specificity of ACE2 binding affinity and RBD expression as predictors of pseudoviral infectivity suggests that changes in ACE2 binding affinity and RBD expression are well-tolerated in many immune-evading mutants, further providing the virus with a range of possibilities for generating mutants that can evade nAbs without compromising infectivity (Fig. 2D).

Taken together, the narrow focus of the immune response on a specific region of the RBD (Fig. 1), the immunodominance of the spike protein (*12*), and the ability of RBD nAb escape mutations to yield viable and infectious viral particles capable of ACE2 binding (Fig. 2) suggest a low evolutionary cost for the virus in generating escape mutants for nAbs.

### Mutant frequency under neutral drift is likely to lead to escape from single and double antibody combinations

Based on this assessment, we used evolutionary theory to predict the frequency of immune escape mutants in the population both before and after the widespread presence of nAbs due to vaccines, passive immunization or natural immunity. Before immunity or antibody prophylaxis is widely established in the population, there is no transmission advantage for viruses with immune escape mutations since most people are equally susceptible to infection from wild-type and mutant SARS-CoV-2. Instead, these mutations may have a small evolutionary fitness cost due to negative effects on ACE2 binding affinity or other factors, similar to the observed fitness cost of drug resistance mutations in HIV (*18*) and consistent with results suggesting that much of the SARS-CoV-2 genome is under weak purifying selection (*19*). Indeed, many point mutations modestly reduce the ability of SARS-CoV-2 to infect cells *in vitro*, which could lead to reduced host-host transmission (*20*). Although these mutants are at a fitness disadvantage compared to the wild-type virus before nAbs are broadly present in the population, they are constantly generated through *de novo* mutation which allows them to exist at nonzero frequencies. However, once nAbs are common in the population, these mutants will have a selective advantage. If they already exist at sufficient frequency in the population, the escape mutants will expand deterministically and lead to widespread SARS-CoV-2 resistance to nAbs.

Using mathematical modeling methods developed to study intrahost evolutionary dynamics during HIV infection, we calculated the expected number of infected individuals whose dominant viral sequence harbors one or more mildly deleterious immune-evading mutations under drift conditions (referred to as “mutants”, see Methods for details) (*21*). Most reported nAbs are susceptible to at least one single-nucleotide change resulting in evasion (*17*), suggesting that a single point mutation may correspond to the evasion of one antibody in a combination. This model predicts the frequency of such mutants using the mutation rate of the virus, the typical fitness cost to the virus from an immune escape mutation, and the total number of infected individuals. We estimated the per base per transmission mutation rate of SARS-CoV-2 from population phylodynamic studies to be between 1×10^−5^ to 1×10^−4^ (Methods) (*2*). Many nAbs are evaded by multiple distinct point mutations, so the per-transmission rate of generating a new mutant that evades a particular neutralizing antibody can be more than an order of magnitude higher than the per base mutation rate (*22*). We investigated a range of infected population sizes (from 5 million to 640 million) and a range of transmission fitness costs for each mutation before the widespread presence of nAbs.

The expected number of SARS-CoV-2-infected individuals whose dominant viral sequence harbors one or two immune escape point mutations is high enough to eventually lead to widespread resistance to nAbs. Over a range of fitness costs and starting from a number of active infections that is lower than the number at present, we predicted that over 10,000 SARS-CoV-2-infected individuals worldwide would harbor a dominant viral sequence capable of evading one antibody (Fig. 3A). This number far exceeds the threshold number of individuals required for natural selection and not neutral genetic drift to drive evolution (dashed lines in Fig. 3). Assuming an immune escape mutant will eventually have a fitness advantage of 0.1, corresponding to approximately 14% of the population receiving an effective prophylactic (Fig. S2), if 10 or more individuals are infected with an immune escape mutant virus, positive selection will allow the mutant to expand and eventually outcompete the wild-type virus (*23*). More than one nucleotide change may be required to confer resistance to an antibody combination if it contains more than one antibody with distinct escape mutation profiles. If a specific two-mutation combination is required for resistance, the expected number of infected individuals harboring a dominant viral sequence capable of evading the antibody combination can be orders of magnitude lower (Fig. 3B). However, substantial double mutant populations (∼hundreds of individuals) are expected if there are more than 50 million active infections worldwide (a plausible count, as of this writing, see Methods for details).

**Figure 3.**
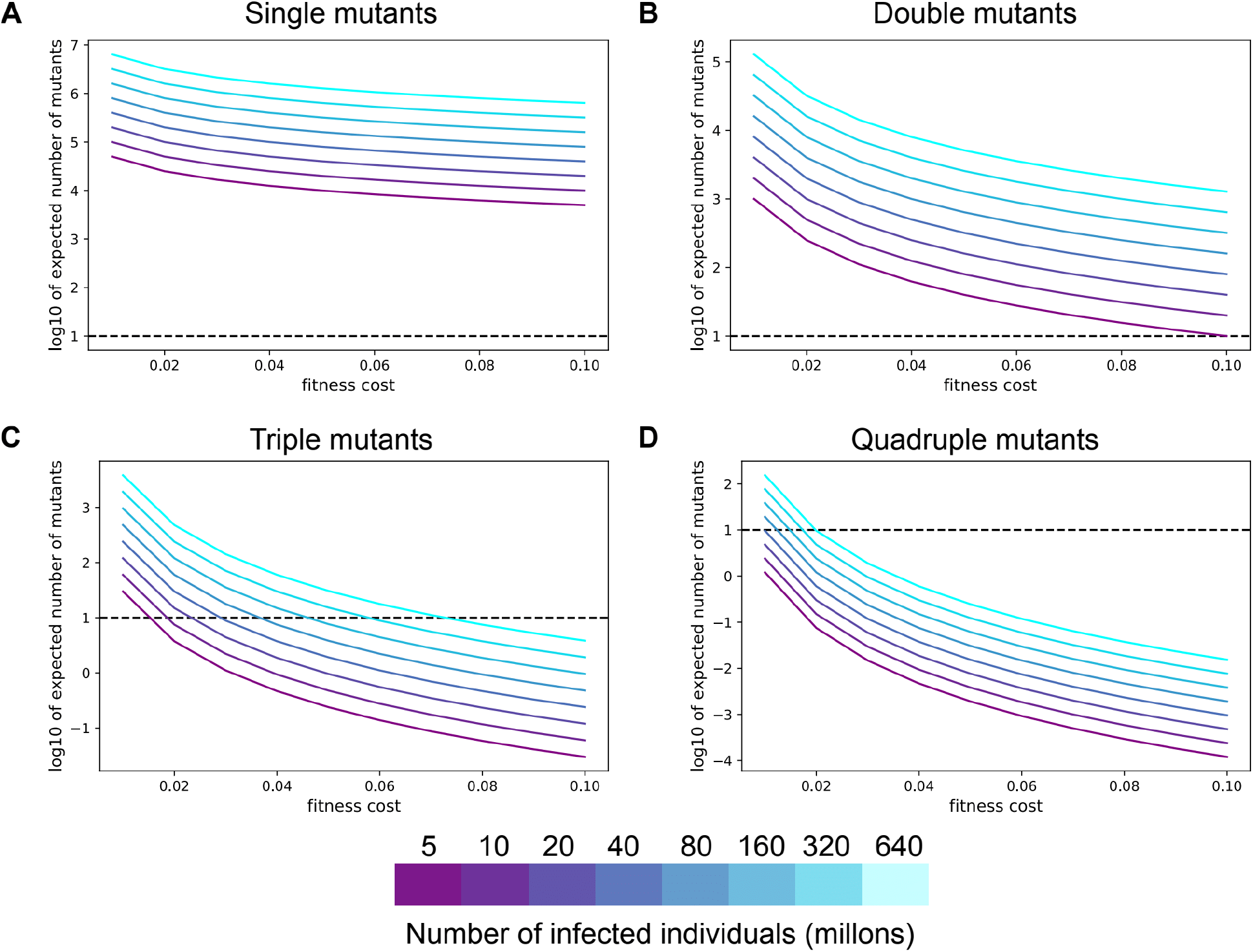
SARS-CoV-2 mutants with one or two mildly deleterious mutations are expected to exist at high numbers. **A-D**. The expected number of individuals infected with a specific single (**A**), double (**B**), triple (**C**), or quadruple (**D**) SARS-CoV-2 mutant viruses at different values of the fitness cost. For all panels, the colors denote the total number of individuals with active SARS-CoV-2 infection globally. The horizontal dashed line is the drift boundary calculated at a fitness benefit of 0.1 for the mutation combination.

If more than two mutations are required for a virus to escape nAbs, it is much less likely that population-level resistance will arise immediately. Each triple mutant is expected to be at appreciable frequencies only when the fitness cost of immune-evading mutations is lower than 0.04 (Fig. 3C). Specific quadruple mutants are not expected to exist at significant frequencies in the population due to standing genetic variation alone for all but the lowest fitness costs (Fig. 3D).

### Single and double resistance mutants are expected to establish quickly under selection

Even if a specific combination of mutations that confers resistance to an antibody combination is not present before the intervention is released, spontaneous mutation and positive selection will eventually lead to expansion of an escape mutant. To estimate how quickly population-level resistance to SARS-CoV-2 antibodies will emerge under natural selection, we modeled the acquisition of multiple mutations over time as a fitness valley-crossing problem (Methods). To acquire a specific combination of mutations that confers therapeutic resistance, the wild-type virus must transit through a valley of intermediate lower-fitness genotypes that have some, but not all, of the mutations required for immune escape. Previous theoretical expressions describing the time required to cross a fitness valley (*24*) were used to estimate the time needed for SARS-CoV-2 to acquire a given combination of one to four mutations (Fig. 4). The time required for establishment of population-level resistance depends on how beneficial resistance is for virus transmission, and this benefit increases as more individuals in the population harbor a specific nAb. For antibodies or antibody combinations capable of being defeated by a single mutation, our modeling predicts the pre-existence of a resistant fraction before deployment of the intervention (Fig. 4A). When examining double mutants, for pandemic sizes of 40 million or more, resistance to a widely deployed nAb combination will occur within months (Fig 4B). However, triple and quadruple mutation combinations will take much longer to establish in the population, even if nAbs are used widely and exert a strong selection pressure for these mutants (Fig. 4C-D). These results hold under a range of intermediate fitness costs for viral mutants that harbor only a subset of the mutations required for escape (Fig. S3).

**Figure 4.**
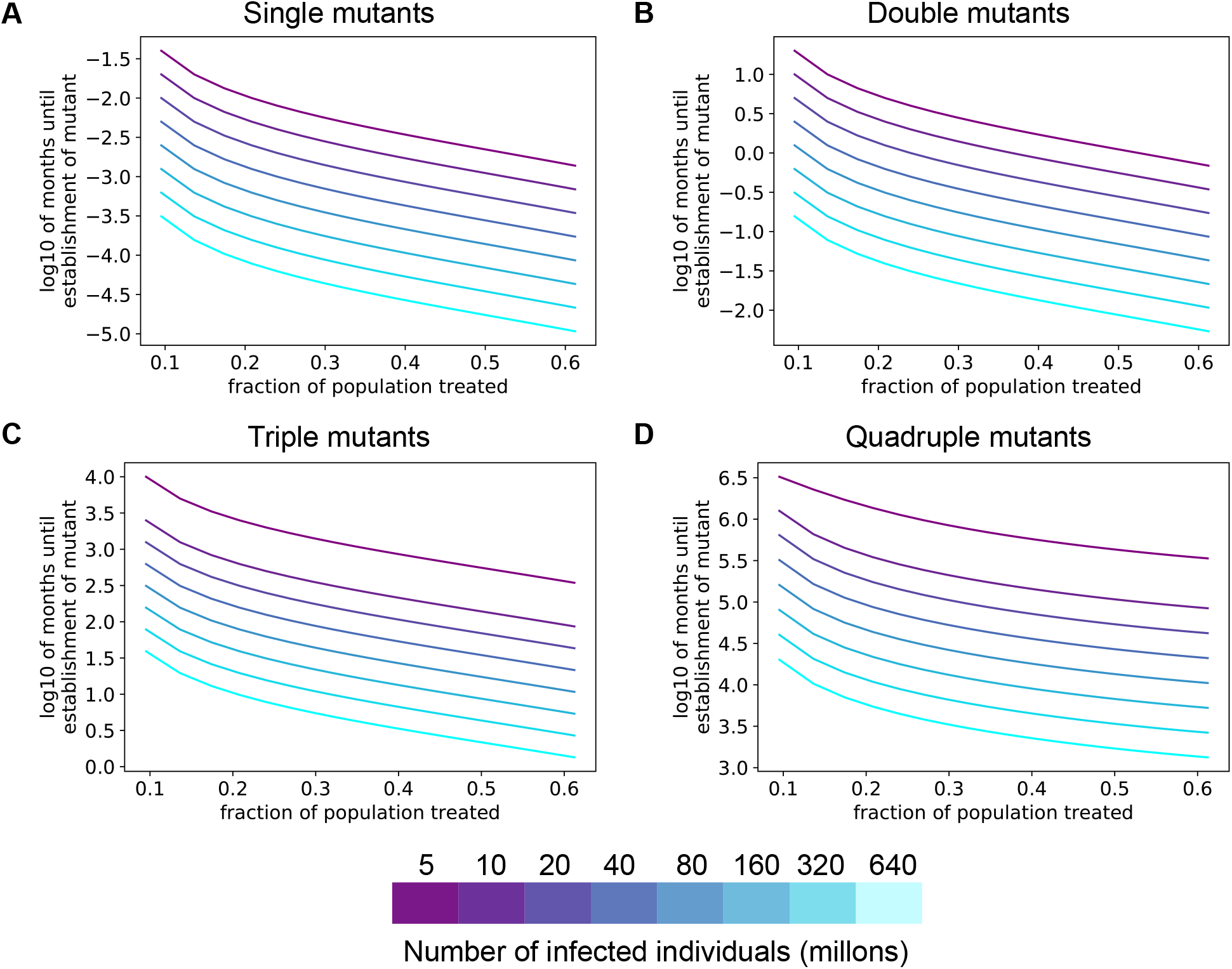
Resistance to single or double antibody combinations will develop quickly under positive selection pressure. **A-D**. Expected time to establishment of a successful single (**A**), double (**B**), triple (**C**), or quadruple (**D**) immune escape mutant assuming a per-site per-transmission mutation rate of 1×10^−4^. The advantageous antibody resistant phenotype is acquired only after a specific combination of 1-4 mutations is present in the same virus. For all panels, the colors denote the total number of individuals with active SARS-CoV-2 infection. The fitness cost for each intermediate mutant is 0.05.

### Population-level resistance occurs more quickly at higher viral mutation rates

The SARS-CoV-2 mutation rate is a key parameter that determines how quickly the virus will acquire resistance to antibody interventions. While we estimated the per transmission rate of generating an antibody escape mutant at 1×10^−4^ (Methods), differences between antibody epitope sizes or changes in the mutation rate of the virus population over time (*25*) could influence this effective mutation rate. Our analysis revealed that many individuals would be infected with single or double SARS-CoV-2 mutants at a range of mutation rates greater than 1×10^−5^ (Fig. 5A-B), while at higher mutation rates even triple and quadruple mutants will occur at sufficient frequencies to quickly establish in the population (Fig. 5C-D). Similarly, we found that resistance to antibody combinations requiring two or fewer mutations for resistance would establish quickly after widespread presence of nAbs (Fig. 5E-F). With a higher mutation rate (1×10^−3^ per transmission), resistance could emerge against even combinations of nAbs that require the acquisition of 4 mutations (Fig. 5H).

**Figure 5.**
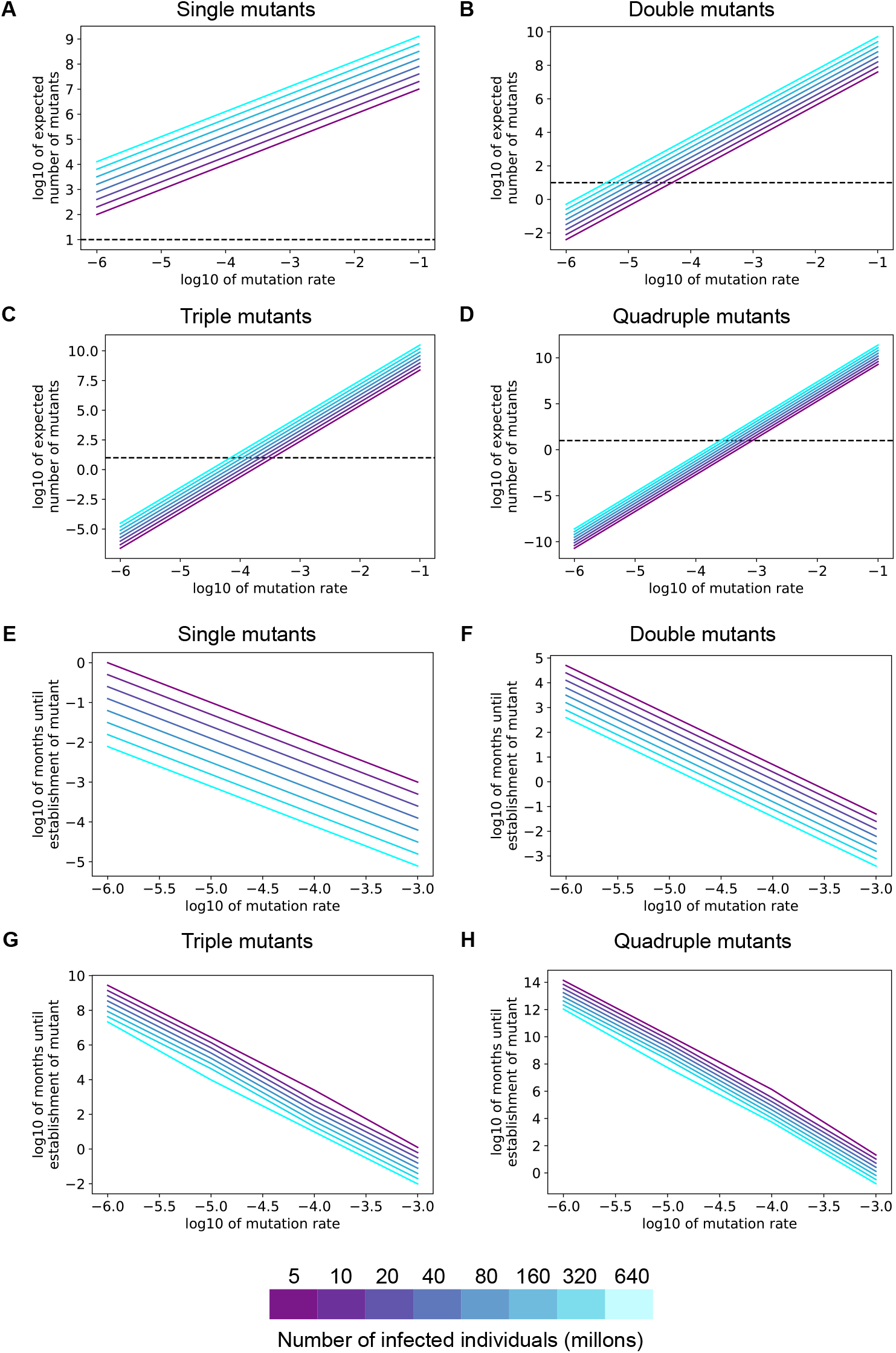
Resistance to single or double antibody combinations will develop quickly across a range of SARS-CoV-2 mutation rates. **A-D**. The expected number of individuals infected with a specific single (**A**), double (**B**), triple (**C**), or quadruple (**D**) SARS-CoV-2 mutant viruses at different values of the per transmission mutation rate. **E-H**. Expected time to establishment of a successful single (**E**), double (**F**), triple (**G**), or quadruple (**H**) immune escape. The fitness benefit of resistance is 0.2, corresponding to 24% of the population receiving an effective prophylactic. For all panels, the colors denote the total number of individuals with active SARS-CoV-2 infection. The fitness cost for each intermediate mutant is 0.05.

In summary, the ability of the virus to readily tolerate nAb resistance mutations means SARS-CoV-2 could quickly evade nAbs due to natural immunity or biomedical interventions.

## Discussion

The spread of SARS-CoV-2 through its newfound human hosts has occurred rapidly, and thus far, in the absence of effective medical countermeasures. Numerous COVID-19 antibody prophylactics and vaccines target the spike protein (*26, 27*), and the immunodominance of the spike RBD in the natural immune response (*12*) implies that even vaccines that use live-attenuated or inactivated SARS-CoV-2 will target the RBD (*28*). Thus, as the next phase of this evolutionary chess game between SARS-CoV-2 and humans unfolds, anticipating the virus’ counter-move to the widespread deployment of spike RBD-targeting nAbs has significant implications for SARS-CoV-2 prophylaxis.

The evolvability of SARS-CoV-2 spike protein RBD in the presence of nAbs depends on two things: the mutation rate in the presence of selection pressure and the mutational tolerance of the spike protein.

The mutation rate of SARS-CoV-2 is in line with that of other single-strand RNA viruses, (*29*), between two-and four-fold lower than that of influenza and HIV (*1, 30*)). Mutation rates for RNA viruses in general are among the highest known (on the order of 10^−6^ per base per viral replication cycle). When compared against many other RNA viruses, such as Hepatitis C, for which evolution has practical clinical consequences, SARS-CoV-2 has a relatively high evolutionary rate (Table S1) (*31, 32*). As mentioned earlier, the currently estimated mutation rate for SARS-CoV-2 is in the context of neutral genetic drift (*19*). Mutation rates themselves are evolvable and increase over time due to natural selection (*33*)-a SARS-CoV-2 RNA dependent RNA polymerase (RdRp) variant that increases the mutation rate by two to five times (eliminating the mutation rate differential with respect to HIV and influenza) has already been identified in some clinical isolates (*25*).

At the same time, the tolerance of spike protein RBD to immune-evading mutations appears to be relatively high. Our analyses demonstrate that experimentally determined immune escape mutations are capable of binding host ACE2, in many cases with little to no loss of affinity relative to wild-type (Fig. 2C). In fact, one prevalent RBD immune-evading mutation (N439K) has been shown experimentally to enhance ACE2 binding and have similar *in vitro* replication fitness to wild-type virus (*34*). Our analyses also demonstrate that compromising spike RBD function (either through loss of ACE2-binding or expression levels) has a weak impact on *in vitro* infectivity (Fig. 2D).

It is likely that standing genetic variation alone has already produced a substantial population of viruses with single and double nucleotide changes that confer nAb resistance. These variants will establish quickly in the population under selection pressure. In fact, there is already precedent for this type of selective sweep, as one such sweep occurred early on in the SARS-CoV-2 pandemic when the D614G mutation rose to nearly 80% frequency in under 6 months (*35*). This mutation confers increased infectivity on the virus (*36*) and was readily generated in sufficient numbers to ensure its expansion. As of this writing, a second selective sweep appears to be underway in Europe, with the 2A.EU1 variant (*37*). Additionally, the recent outbreak in minks of a variant with a combination of mutations that reduce antibody binding suggests that even variants with multiple mutations can be generated and are viable (*38*). Currently, most of the SARS-CoV-2 genome is not under positive selection (*19*), but if nAbs are widely present in the population, mutations that confer resistance via immune evasion will expand rapidly under positive selection pressure.

The current consensus in the scientific literature is that SARS-CoV-2 mutations and evolution are not likely to change the course of this pandemic (*30, 39, 40*). One line of reasoning is that generating viral mutants with higher fitness is difficult because very few individual nucleotide changes are likely to increase viral replication, transmission, or infectivity. However, this argument confuses the rate of evolution in the absence of selection pressure (neutral genetic drift) with the rate of evolution under natural selection. To this end, our analysis suggests that mutations capable of evading nAbs with limited impact on viral fitness are already broadly present (Fig. 2). The N439K immune-evading spike mutant was reported to maintain replicative fitness in patients (*34*), supporting this idea. A second line of reasoning is that because the SARS-CoV-2 mutation rate is relatively low, evolution will be too slow to be consequential. However, the reported mutation rate under drift is not particularly low, is likely to increase under selection pressure, and is high enough at present to generate single and double immune-evading mutants at a concerning frequency.

Going forward, our work suggests strategies for designing SARS-CoV-2 prophylactics that are more resistant to viral evolution. First, nAbs should be used in combinations, preferably targeting more than two non-overlapping epitopes. Strategies for the design of prophylactic antibodies and vaccines should involve combining nAbs that bind to non-overlapping escape mutant regions, including those from smaller, distinct clusters outside the RBD. Alternatively, if antibodies from the same cluster are used, escape mutants must be carefully characterized to ensure they do not overlap (*22*). Second, the evolutionary pressure on the virus will determine the speed at which resistance to nAbs emerges. The more ubiquitous a nAb is, and the more effective it is, the faster it will generate resistance (Fig. 4). This is a potential weakness of focusing on only one or two vaccines for global deployment. The effectiveness of nAb-based interventions for disease control will depend on how many different interventions are deployed, how many mutations are required to evade each intervention, and the extent to which their escape mutations overlap. Finally, the overall size of the pandemic in terms of number of active infections will play a significant role in whether the virus can be brought under control with nAbs prophylactics or vaccines.

Additionally, SARS-CoV-2 antigen tests based on nAbs (*41, 42*) could impose a selective pressure on the targeted epitope(s). Our work suggests that these tests should be based on multiple antibodies targeting distinct epitopes.

The evolvability of SARS-CoV-2 in response to selection pressure will determine the ultimate tractability of our efforts at disease control. Our findings are relevant to the viability of any prophylactic modality which can be evaded by a series of single-nucleotide mutations. Similar questions can and should be asked of the viral capacity to evade the cellular adaptive immune response and the ability of other viral proteins to escape nAbs. However, additional investigation into the diversity of the nAb response to vaccines and the spatial and temporal dynamics of the host immune response is necessary to fully understand the risk of resistance to vaccines. Additionally, reports that SARS-CoV-2 mutates sufficiently rapidly within each patient to form a quasispecies suggests that genetic diversity within hosts may be sufficient to lead to acquired resistance to therapeutic nAbs (*43*).

## Data Availability

All data used in the manuscript is publicly available, through the GISAID database or the cited references.

## Acknowledgements

### Funding

D.V.E. acknowledges funding from the NSF-Simons Center for Mathematical and Statistical Analysis of Biology at Harvard, award number #1764269, and the Harvard Quantitative Biology Initiative. A. N. acknowledges funding from the National Science Foundation Graduate Research Fellowship Program under Grant No. DGE-1762114. Any opinions, findings, and conclusions or recommendations expressed in this material are those of the author(s) and do not necessarily reflect the views of the National Science Foundation.

### Author contributions

A.C. conceived of the study. A.C., D.V.E., M.S., and A.N. designed the study approach. D.V.E., M.S., A.N., and U.T. implemented the analyses and created the figures. A.C., D.V.E., M.S., and A.N. wrote the initial draft of the manuscript, which was edited by all authors. All authors read and approved the final manuscript. D.J.-M. and A.C. supervised the study, with input and feedback on specific topics from M.H., J.M.C., B.Z., B.L.P., and M.R.

### Competing interests

A.C., M.S., and U.T. are employees and shareholders of Fractal Therapeutics. D.V.E., A.N., B.Z., and D.J.-M. are shareholders of Fractal Therapeutics.

### Data and materials availability

Scripts for running and plotting the evolutionary models are available at on GitHub at https://github.com/dvanegeren/covid-nAb-escape.

## Materials and Methods

### Compilation of published neutralizing antibody epitopes

The authors performed a comprehensive search of all entries in the Protein Data Bank (PDB) (*44*) as of September 1^st^, 2020 which matched the criteria “Source Organism Taxonomy Name equals SARS-2 AND Source Organism Taxonomy Name equals Homo sapiens”. Structures were included if there were patient-derived nAbs present and the authors reported the binding residues. Structures were excluded if they were not patient-derived, if they were not nAbs, or if the authors did not report the binding epitope because the resolution was too low to identify it precisely. Additionally, other epitopes that met the inclusion criteria but were not found in the PDB were included on an ad hoc basis. The structures used are listed in Table S3.

After the search was complete, it was determined that there were too few epitopes reported outside of the RBD to attempt clustering in those residues. Thus, the clustering analysis was limited to the RBD. The RBD was defined as in (*17*).

### Clustering of antibody epitopes

The antibodies were clustered using the Density-Based Spatial Clustering of Applications with Noise (DBSCAN) algorithm (*45*). The distance function used to compare two epitopes was the median distance between residues in the epitopes. The maximum distance between two epitopes in one cluster was set at 30 residues. The minimum number of epitopes per cluster was set at 2. Epitopes classified as noise were assigned to their own clusters. These parameters were set based on a subset of 9 antibodies and used for all subsequent clustering. They were also tested on larger sets that included non-neutralizing antibodies (data not shown). Clustering was done using Python version 3.7 with the scikit-learn package (*46*).

### ACE2 binding affinity and RBD predictivity for pseudoviral infectivity

Tolerability of SARS-CoV-2 to changes in ACE2 binding affinity or RBD expression were examined based on a receiver operating characteristic (ROC) analysis. Based on publications measuring the impact of 44 spike RBD mutations on the *in vitro* infectivity of SARS-CoV-2 pseudovirus, we examined the predictivity of changes in RBD binding to ACE2 or expression according to (*17*) for greater than 10% loss of infectivity as measured by luciferase reporter pseudoviral assay (*15*). These mutants were either observed circulating in the population in GISAID (*20*) or experimentally identified to confer escape from nAbs (*17, 47*). Predictivity of ACE2 binding affinity or RBD expression for *in vitro* infectivity was evaluated based on ability to predict a 90% or greater infectivity relative to the WT strain.

### Expected number of mutants without positive selection

To investigate the emergence of SARS-CoV-2 resistance to nAbs, we modeled virus transmission dynamics using a modified deterministic susceptible-infectious-recovered (SIR) model with mutation. The simplest version of this model includes two viral genotypes, wild-type (WT) and mutant viruses with a specific single nucleotide change. At the population level there are four different compartments: susceptible individuals (*S*), individuals infected with WT virus (*I*_0_), infected with mutant virus (*I*_1_), and recovered or resistant to infection from WT virus (*R*). The number of individuals in each compartment obey the following differential equations:

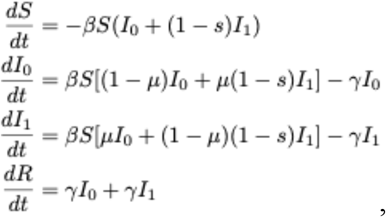

where *β* is the transmission rate of WT virus, *μ* is the per transmission probability of mutation at a specific site, *s* is the fitness cost to transmission of the mutation, and *γ* is the recovery rate from the infection. The infected compartments of this SIR model have the same mathematical description as the virus-infected cells in the intrahost virus dynamics model presented by Ribeiro and co-workers (*21*), assuming the birth and death rates of uninfected cells in the intrahost model is negligible. We also assume that the frequency of recovered individuals in the population is small enough so *S* and *R* can be treated as constant. Therefore, for the frequency of virus mutants present in the population at steady state before establishment of widespread immunity or vaccination, we used the frequency of individuals infected with a virus with a single mutation *f*_1_ = *μ*/*s*, which agrees with the expected frequency of single mutants under mutation-selection equilibrium (*24*). As given in (*21*), the frequency of viral mutants with *k* mutations is

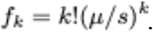

### Expected time to development of population-level resistance under positive selection

If a substantial fraction of the population is immune to the WT virus (either due to lasting immunity after recovering from the infection, vaccination, or administration of therapeutic antibodies), viruses with antibody escape mutations will have a fitness advantage over the WT virus. This advantage comes from the ability of the mutant virus to infect individuals who are immune to the WT virus and depends on the fraction of the population who are immune. If a single mutant has the ability to infect recovered/resistant individuals, the SIR model equations for infected individuals change to

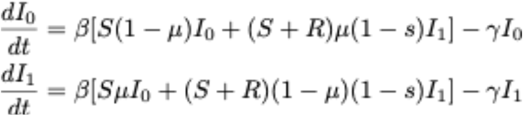

and the effective transmission rate for the mutant virus is given by 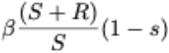.

The selective advantage *w* of the mutant virus is therefore 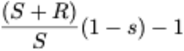.

In order for a virus with one or more mutations that confer immune escape to expand deterministically due to positive selection and establish in the population, the variant must first be created through mutation of a single virion. Then, the mutant virus must infect enough individuals so that it is unlikely to go extinct due to stochastic drift. Assuming the total number of infected individuals *N* is constant, if a single mutation is sufficient to lead to immune escape, the time needed to establish an immune escape mutant is exponentially distributed with expected time 1/*Nμw* generations (*24*).

To estimate the time needed for establishment of double-, triple-, or higher-order mutants that confer immune escape, we adapted previously reported work on the dynamics of asexual populations crossing fitness valleys (*24*). If *k* mutations are required for immune escape and all intermediates with less than *k* mutations have fitness cost *s*, the time *τ*_*k*_ to establishment of the *k*-mutant was approximated as

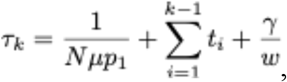

where *γ* is Euler’s constant, 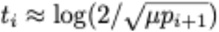, and the probability *p*_*i*_ of an *i*-mutant to be successful was approximated as

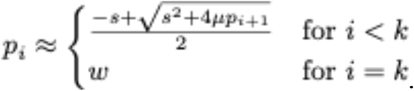

This approximation holds for intermediate population sizes (*Nμ* < 1). Following the argument for large populations in (*24*), when *Nμ* > 1 we treated mutants with few mutations deterministically. We again used the results in (*21*) to estimate the frequency of *i*-mutants at steady state under mutation-selection equilibrium *f*_*i*_ for *i* <= *k*. The minimum value of *i* for which the estimated i-mutant population size *Nf*_*i*_ was < 1/*μ* was taken as the mutant with the most mutations that could be treated deterministically as a constant-sized population. Denoting this number of mutations as j, the modified expression for the expected time to escape for large population sizes is

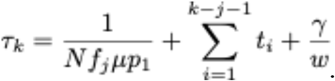

### Evolutionary model parameter value selection

The SARS-CoV-2 infection length was set to 2 weeks, based on published estimates of infectious period length (*48*).

The effective rate of acquiring nucleotide substitutions that escape a given nAb was estimated to be 1×10^−4^ per transmission. The overall single nucleotide substitution rate has been estimated at approximately 1×10^−3^ per site per year from multiple phylogenetic analyses of global SARS-CoV-2 genomic sequences (*2*), which is a 3.8×10^−5^ per site per transmission mutation rate assuming a 2 week infection generation time. However, since multiple different single nucleotide mutations have been shown to confer resistance to many nAbs (*22*), we estimated the effective mutation rate (defined as the per transmission rate of producing a mutation that generates resistance to a particular nAb) to be 2-3x higher than the rate of producing a particular nucleotide substitution. The effect of changing the mutation rate on the mutant frequency and escape time estimates is shown in Fig. 5.

We assumed that, without selective pressure imposed by deployment of an intervention, mutant virus is less fit than wild-type virus and is not transmitted as effectively. A similar fitness cost is assumed to apply to mutant intermediates that only harbor a subset of mutations required for escape from an antibody combination. This fitness cost to viral transmission is difficult to directly measure, so we used a range of fitness cost from 0.01 to 0.1, corresponding to a 1-10% reduction in transmission rate for mutant viruses. These fitness costs are of similar magnitude to those measured for HIV drug resistance mutations in treatment-naïve patients (*18*) and are broadly justified by the limited impact of spike RBD mutations on ACE2 binding and the limited ability of ACE2 binding and expression to predict infectivity (Fig. 2).

We estimated the total number of individuals infected with SARS-CoV-2 using the number of diagnosed cases. As of 11/8/20, the number of active diagnosed cases worldwide is 14 million (*49*), and the number of infections is expected to be 5-10 times the number of diagnosed cases, as determined by modeling (*50*) and seroprevalence studies (*51*).

