## Supplementary material for "Risk of evolutionary escape from neutralizing antibodies targeting SARS-CoV-2 spike protein": Tables S1-S3 and Figures S1-S3

**Table S1.** Evolutionary rates of pathogenic RNA viruses

| Virus | Type | Evolutionary rate ($10^{-3}$ sub/site/year) | Reference |
|---|---|---|---|
| HIV | Retrovirus | 2.02 – 16.8 | (*1*) |
| Poliovirus | +ssRNA | 10.3 | (*2*) |
| Influenza A | -ssRNA | 1.43 - 1.16 | (*3*) |
| SARS-CoV | +ssRNA | 7.8 | (*4*) |
| SARS-CoV-2 | +ssRNA | 0.8 - 6.58 | (*5*) |
| Rotavirus | dsRNA | 0.73 | (*6*) |
| Hepatitis C virus | +ssRNA | 0.48 – 0.91 | (*7*) |
| MERS-CoV | +ssRNA | 0.24 | (*4*) |

**Table S2.** Antibody escape mutants with highest ACE2 binding affinities.

| Antibody | % Change in Binding Affinity for Optimal Escape Mutant |
|---|---|
| S309 | 120% |
| EY6A | 155% |
| S2A4 | 120% |
| S304 | 155% |
| S2M11 | 117% |
| B38 | 200% |
| C105 | 178% |
| P2B-2F6 | 141% |
| CB6 | 151% |
| CC12.1 | 200% |
| CC12.3 | 200% |
| CV30 | 200% |
| Fab 2-4 | 178% |
| COVA2-04 | 200% |
| COVA2-39 | 200% |
| S2H13 | 200% |
| S2H14 | 200% |
| S2E12 | 123% |
| REGN10933 | 178% |
| REGN10987 | 200% |

**Table S3**. Antibody structures used.

| Antibody Name | Structure Type | Source |
|---|---|---|
| S309 | Cryo-electron Microscopy and Crystal Structure | (*1*) |
| EY6A | Cryo-electron Microscopy and Crystal Structure | (*2*) |
| S2A4 | Cryo-electron Microscopy and Crystal Structure | (*3*) |
| S304 | Cryo-electron Microscopy and Crystal Structure | (*3*) |
| S2M11 | Cryo-electron Microscopy | (*4*) |
| B38 | Crystal Structure | (*5*) |
| C105 | Cryo-electron Microscopy of complex. Crystal structure of antibody. | (*6*) |
| P2B-2F6 | Crystal Structure | (*7*) |
| CB6 | Crystal Structure | (*8*) |
| CC12.1 | Crystal Structure | (*9*) |
| CC12.3 | Crystal Structure | (*9*) |
| CV30 | Crystal Structure | (*10*) |
| Fab 2-4 | Cryo-electron Microscopy | (*11*) |
| COVA2-04 | Crystal Structure | (*12*) |
| COVA2-39 | Crystal Structure | (*12*) |
| S2H13 | Cryo-electron Microscopy | (*3*) |
| S2H14 | Cryo-electron Microscopy and Crystal Structure | (*3*) |
| S2E12 | Cryo-electron Microscopy of complex. Crystal structure of antibody. | (*4*) |

1. D. Pinto, Y.-J. Park, M. Beltramello, A. C. Walls, M. A. Tortorici, S. Bianchi, S. Jaconi, K. Culap, F. Zatta, A. De Marco, A. Peter, B. Guarino, R. Spreafico, E. Cameroni, J. B. Case, R. E. Chen, C. Havenar-Daughton, G. Snell, A. Telenti, H. W. Virgin, A. Lanzavecchia, M. S. Diamond, K. Fink, D. Veesler, D. Corti, Cross-neutralization of SARS-CoV-2 by a human monoclonal SARS-CoV antibody. *Nature*. **583**, 290–295 (2020).

2. D. Zhou, H. M. E. Duyvesteyn, C.-P. Chen, C.-G. Huang, T.-H. Chen, S.-R. Shih, Y.-C. Lin, C.-Y. Cheng, S.-H. Cheng, Y.-C. Huang, T.-Y. Lin, C. Ma, J. Huo, L. Carrique, T. Malinauskas, R. R. Ruza, P. N. M. Shah, T. K. Tan, P. Rijal, R. F. Donat, K. Godwin, K. R. Buttigieg, J. A. Tree, J. Radecke, N. G. Paterson, P. Supasa, J. Mongkolsapaya, G. R. Screaton, M. W. Carroll, J. Gilbert-Jaramillo, M. L. Knight, W. James, R. J. Owens, J. H. Naismith, A. R. Townsend, E. E. Fry, Y. Zhao, J. Ren, D. I. Stuart, K.-Y. A. Huang, Structural basis for the neutralization of SARS-CoV-2 by an antibody from a convalescent patient. *Nature Structural & Molecular Biology*. **27**, 950–958 (2020).

3. L. Piccoli, Y.-J. Park, M. A. Tortorici, N. Czudnochowski, A. C. Walls, M. Beltramello, C. Silacci-Fregni, D. Pinto, L. E. Rosen, J. E. Bowen, O. J. Acton, S. Jaconi, B. Guarino, A.

Minola, F. Zatta, N. Sprugasci, J. Bassi, A. Peter, A. D. Marco, J. C. Nix, F. Mele, S. Jovic, B. F. Rodriguez, S. V. Gupta, F. Jin, G. Piumatti, G. L. Presti, A. F. Pellanda, M. Biggiogero, M. Tarkowski, M. S. Pizzuto, E. Cameroni, C. Havenar-Daughton, M. Smithey, D. Hong, V. Lepori, E. Albanese, A. Ceschi, E. Bernasconi, L. Elzi, P. Ferrari, C. Garzoni, A. Riva, G. Snell, F. Sallusto, K. Fink, H. W. Virgin, A. Lanzavecchia, D. Corti, D. Veesler, Mapping Neutralizing and Immunodominant Sites on the SARS-CoV-2 Spike Receptor-Binding Domain by Structure-Guided High-Resolution Serology. *Cell*. **183**, 1024-1042.e21 (2020).

4. M. A. Tortorici, M. Beltramello, F. A. Lempp, D. Pinto, H. V. Dang, L. E. Rosen, M. McCallum, J. Bowen, A. Minola, S. Jaconi, F. Zatta, A. D. Marco, B. Guarino, S. Bianchi, E. J. Lauron, H. Tucker, J. Zhou, A. Peter, C. Havenar-Daughton, J. A. Wojcechowskyj, J. B. Case, R. E. Chen, H. Kaiser, M. Montiel-Ruiz, M. Meury, N. Czudnochowski, R. Spreafico, J. Dillen, C. Ng, N. Sprugasci, K. Culap, F. Benigni, R. Abdelnabi, S.-Y. C. Foo, M. A. Schmid, E. Cameroni, A. Riva, A. Gabrieli, M. Galli, M. S. Pizzuto, J. Neyts, M. S. Diamond, H. W. Virgin, G. Snell, D. Corti, K. Fink, D. Veesler, Ultrapotent human antibodies protect against SARS-CoV-2 challenge via multiple mechanisms. *Science* (2020), doi:10.1126/science.abe3354.

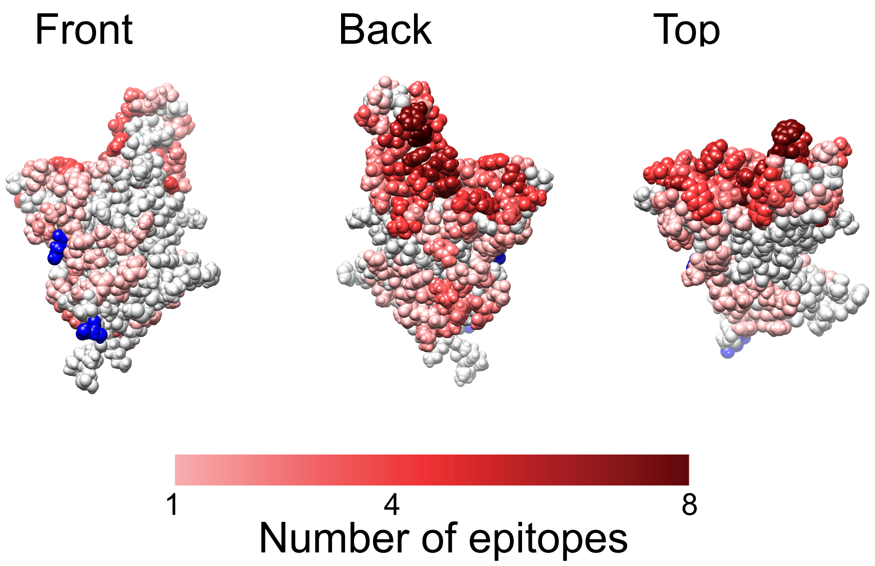


**Figure S1.** Glycosylation in SARS-CoV-2 spike protein RBD. Glycosylated residues are marked in blue, while the remainder of residues are colored by the number of epitopes that contain the residue (red color bar).

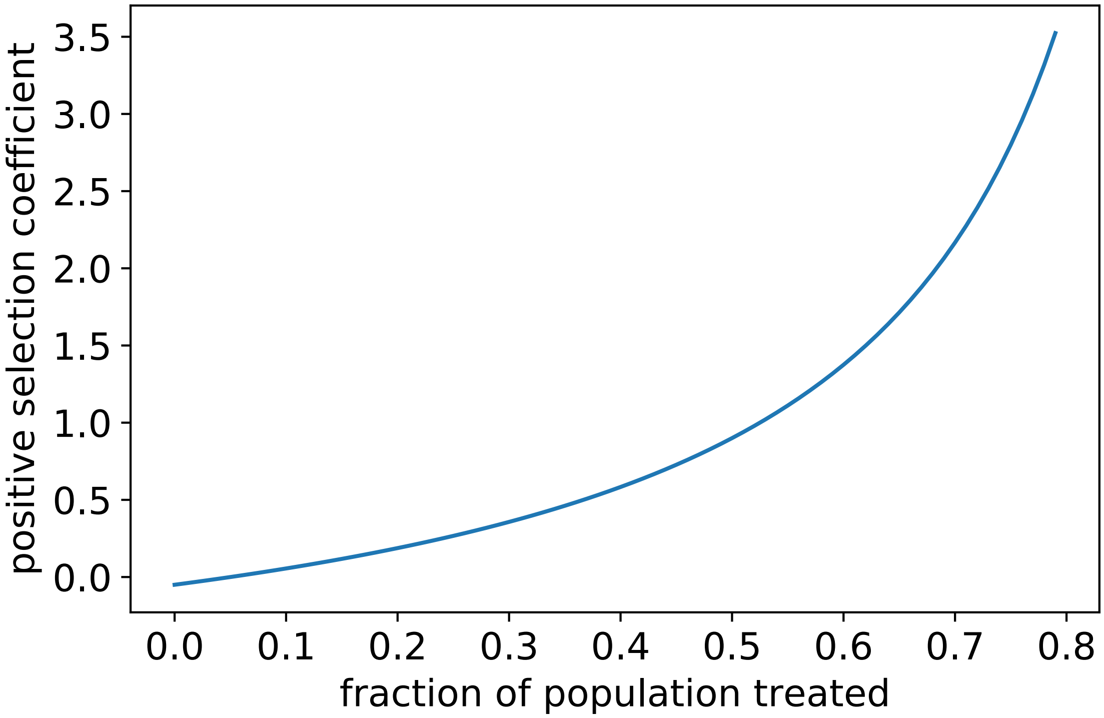


**Figure S2.** Relationship between the fraction of the population that receive a prophylactic that is completely effective in preventing infection from wild-type virus and the strength of selection for an escape mutant.

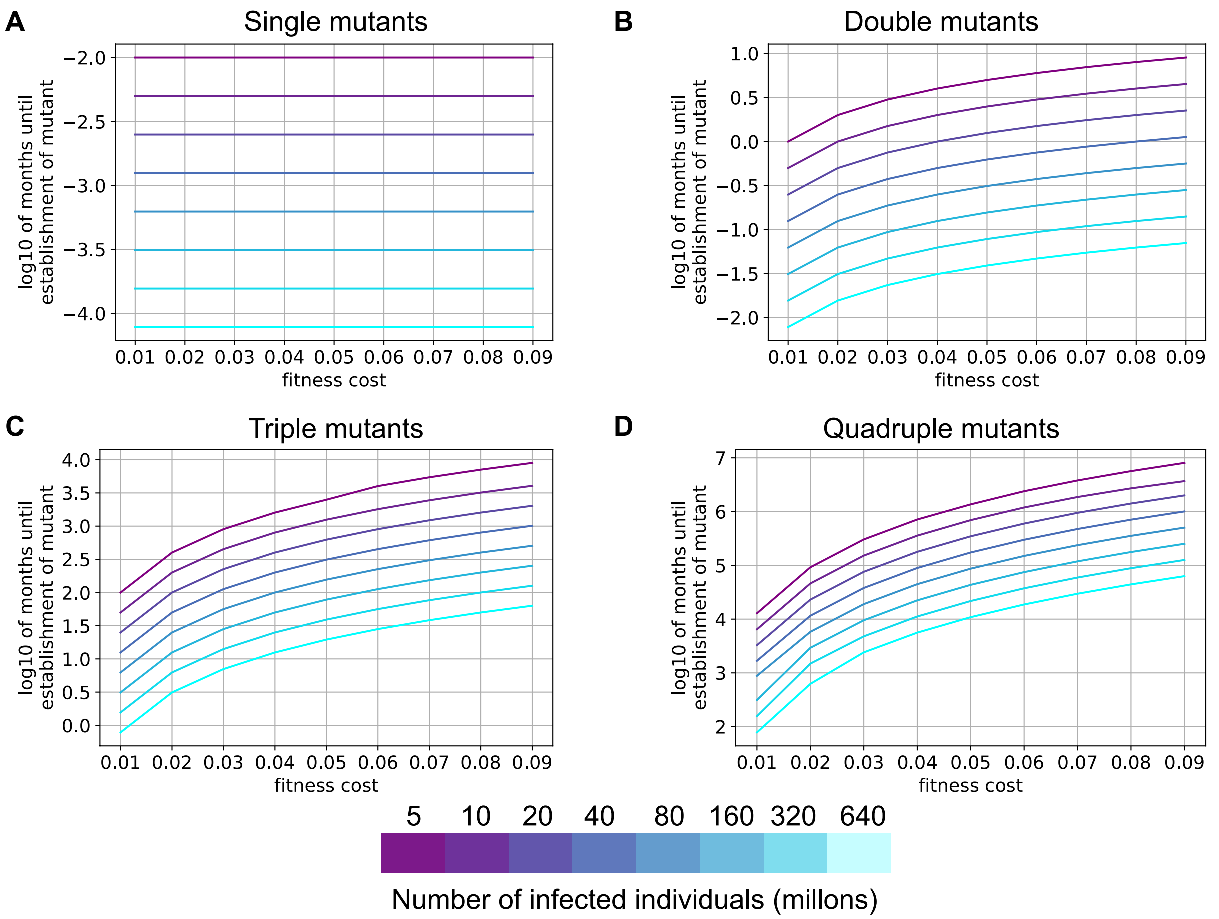


**Figure S3.** Time required to establish a resistant viral single (**A**), double (**B**), triple (**C**), or quadruple (**D**) mutant with different fitness costs for intermediate mutants. In our model, viral variants with some, but not all, mutations required for resistance to an antibody intervention have a fitness cost (ranging from 1-9% less infectious). Increasing the fitness cost of these intermediates prolongs the time required for a resistant variant with a specific combination of 2-4 mutations (**B-D**) to establish in the population.
